# PROSPECTIVE SCREENING OF WILSON DISEASE IN PRIMARY SCHOOL CHILDREN USING SPOT URINE: AN UNFORESEEN SUCCESS IN CASE DIAGNOSIS IN A PILOT STUDY

**DOI:** 10.1101/2025.08.20.25333881

**Authors:** Anne Mei-Kwun Kwok, Joannie Hui, Iris H S Chan, N H Chiang, Toby C H Chan, L Y Hung, Timothy H T Cheng, Matthew Yeung, Xingyan Wang, KM Belaramani, Felix S D Yam, Yuk Him Tam, Chloe Miu Mak, Albert Martin Li, C W Fung, Nelson L S Tang

**Affiliations:** Department of Paediatrics and Adolescent Medicine, Hong Kong Children’s Hospital, Hong Kong SAR; CUHK Medical Centre, Shatin, Hong Kong SAR; Department of Chemical Pathology, Prince of Wales Hospital, Hong Kong SAR; Department of Chemical Pathology, Faculty of Medicine, The Chinese University of Hong Kong, Hong Kong SAR; Department of Pathology, Hong Kong Children’s Hospital, Hong Kong SAR; Department of Surgery, Hong Kong Children’s Hospital, Hong Kong SAR; Department of Paediatrics, Faculty of Medicine, The Chinese University of Hong Kong, Hong Kong SAR; Li Ka Shing Institute of Health Sciences, Faculty of Medicine, The Chinese University of Hong Kong, Hong Kong SAR

**Keywords:** Wilson disease, population screening, spot urine copper, pre-symptomatic diagnosis, zinc therapy

## Abstract

**Background:** Wilson disease (WD) is a rare but commonly under-diagnosed inherited metabolic disease. Patients who are diagnosed early before disease onset (pre-symptomatic WD) have a good response to treatment. For universal screening in children, spot urine tests are more feasible than 24-hour urine collection. We previously established reference ranges for spot urine copper excretion indices. Here, we evaluated their screening performance in a prospective cohort of school children.

**Methods:** Two samples of spot urine and one 24-hour urine were collected from 193 healthy Hong Kong children aged 4-11 years. Urine copper was measured by Inductively coupled plasma mass spectrometry (ICP-MS). Previously established screening cut-off values were evaluated: spot urine copper ≥0.5 µmol/L, copper to osmolality ratio ≥0.00085 μmol/mOsm, and copper to creatinine ratio ≥0.1 µmol/mmol together with a new step cut-off according to urine osmolality. Children whose urine sample exceeded any one cut-off value were called back for blood ceruloplasmin and copper as second-tier tests together with diagnostic sequencing of the *ATP7B* gene.

**Results:** 10 children (5%) had second-tier testing. Two children had very low ceruloplasmin levels and were genetically confirmed to have WD. Both were completely asymptomatic at diagnosis. Spot urine copper concentration ≥0.5 µmol/L showed the best screening performance with excellent sensitivity. The strong correlation between spot urine copper concentration and 24-hour urine copper excretion (R²=0.83, p<0.01) provided the basis for screening WD by spot urine copper. In addition, one carrier was found among the call-back group who had normal plasma ceruloplasmin.

**Conclusions:** In this small prospective screening cohort, 2 WD patients and 1 carrier were diagnosed. Spot urine copper is a useful biomarker for universal WD screening in school children, which may improve disease outcome and fundamentally change the natural history of WD by enabling early detection and therapy before symptom onset.

## INTRODUCTION

Wilson disease (WD, MIM# 277900) is an inherited disorder of copper metabolism (Roberts and Schilsky 2023). It is an autosomal recessive disease caused by mutations in the *ATP7B* gene. A defective ATP7B protein impairs copper incorporation into ceruloplasmin and biliary excretion of copper, resulting in copper accumulation and toxicity in the liver and brain (Członkowska et al. 2018). The incidence of WD varies among populations, with high prevalence found in Southern China and some Mediterranean areas (J. Wang and Abuduxikuer 2019; Sandahl et al. 2020; Loudianos et al. 2024). We characterised the prevalence of various inherited metabolic diseases (IMDs) in the Southern Chinese population, and WD is one of the more common IMDs (Hui et al. 2014; 2013; Mak et al. 2008). Prevalence of WD in Hong Kong may be as high as 1 in 5,400 (Mak et al 2008). The data confirms that WD is an important disease and there is a need for early diagnosis and treatment.

It may be challenging to establish the diagnosis as the presenting symptoms of WD vary. Main clinical symptoms can be broadly categorised into hepatic, neurological or psychiatric symptoms with variable age of onset and severity (European Association for the Study of the Liver 2025). Over the past decade, cascade screening of family members of probands and a high level of clinical suspicion had led to an increasing number of WD patients being diagnosed at a pre-symptomatic stage (Członkowska et al. 2018; Dhawan et al. 2005; Socha et al. 2018; Nicastro et al. 2010; Loudianos et al. 2024). Since copper accumulation and tissue damage take time to develop, initiating treatment before symptoms onset could result in a better prognosis. Our case series and other larger-scale follow-up studies of pre-symptomatic WD confirmed that zinc therapy is highly effective, preventing clinical symptoms of copper toxicity in many pre-symptomatic patients (Hui et al. 2013; Eda et al. 2018; Nakayama et al. 2008; Ranucci et al. 2014; Roberts and Schilsky 2023). Clinical disease onset is rare before puberty (Li et al. 2024; Loudianos et al. 2024). This roughly ten-year pre-symptomatic window offers a valuable opportunity for universal WD screening and early treatment.

Universal screening for WD requires a biomarker that is both sensitive and specific. Previously, researchers attempting to screen for WD using blood biomarkers (like ceruloplasmin and its isoforms) during the newborn or infancy period reported only limited success (Hahn 2014; Roberts and Schilsky 2023; Socha et al. 2018). Recently, a sensitive assay measuring ATP7B peptides, based on immunoaffinity enrichment combined with selected reaction monitoring mass spectrometry (immune-SRM), was developed and evaluated with newborn screening (NBS) blood spot samples (Collins et al. 2021; Klippel et al. 2025). However, the false-negative rate was up to 7.9% (17 out of 216 WD patients) in the evaluation (Collins et al. 2021). Moreover, intervention is unikely to be provided during infancy before the patients are exposed to significant dietary copper intake. NBS results may also cause stress in the families, especially when no treatment or intervention is provided at the time of diagnosis. Therefore, alternative screening strategies and the optimal screening age need to be explored (Chanpong and Dhawan 2022). Other new blood/serum based tests have been developed recently including accurate non-ceruloplasmin copper (Sandahl et al. 2025) and speciation non-ceruloplasmin copper (Schilsky et al. 2022) but they are not widely accessible. Mutation-based screening is also feasible, but it also comes with its problem such as difficulties in predicting functional consequences for new variants (e.g. VUS) identified (Espinós and Ferenci 2020).

Increased urinary copper excretion is a functional defect and hallmark feature of WD. While classical teaching suggested that urine copper measurements might not be a reliable biomarker during acute decompensation in WD patients, this may not apply in the context of screening (Perman et al. 1979; Martins da Costa et al. 1992). It may be a useful biomarker for WD screening since the target is healthy children in the setting of universal screening. 24-hour urine copper excretion is a key diagnostic feature, it is not practical to collect 24-hour urine samples in young children for screening. On the other hand, spot urine collection is more practical and may serve as a proxy index of daily excretion. Application of such proxy indices has been reported for various spot urine analytes (Sieniawska et al. 2012; B. Wang et al. 2015; Yeh et al. 2015). We have studied the use of short-period urine collection in the evaluation of the body status of Magnesium(N. L. S. Tang et al. 2000). Other groups carried out pilot studies in the use of spot urine in the assessment of body status or dietary intake of sodium (Kelly et al. 2015; McLean et al. 2014; Subramanian et al. 2013), magnesium and other analytes (Sieniawska et al. 2012; Touitou et al. 2010). Furthermore, a previous investigation of spot urine copper excretion indices had been carried out by our team and others (N. L. Tang et al. 2020; J.-S. Wang et al. 2010). In a recent review of WD biomarkers, it was suggested that further studies are needed to evaluate the potential use of spot urine copper indices in WD (Chanpong and Dhawan 2022; Armer and De Goede 2017).

As there was little information about the normal level of urinary copper excretion in healthy children, we carried out a retrospective study using archival samples of spot urine collected from newly diagnosed pre-symptomatic WD children and healthy children (N. L. Tang et al. 2020). We found that the difference in spot urine copper indices (e.g. spot urine copper concentration) between WD and healthy children was more than 2-fold in magnitude and potential cut-off values of spot urine indices have been proposed for use in universal screening of pre-symptomatic WD. In this study, these cut-off values were evaluated in a new prospective cohort of children between 4 to 11 years old for validation. This study led to the diagnosis of 2 pre-symptomatic WD children and 1 carrier. This finding provides compelling evidence for the feasibility of non-invasive universal screening for WD among school children using two spot urine samples.

## METHODS

### Study Design and Participants

This was a prospective screening study targeting Hong Kong school children, aged 4-11 years. Inclusion criteria were healthy children with no known history of liver, kidney, neurological, or trace element metabolism disorders. Children with acute illness at the time of sample collection were excluded. The study has been approved by the Institutional Review Board of the Hong Kong Children’s Hospital and the Joint CUHK-NTEC Clinical Research Ethics Committee. Informed consent was obtained from parents or guardians of all participants.

### Sample Collection and Handling

Recruitment of healthy school children aged 4-11 years to participate in the study was conducted from August 2022 to September 2024. Recruitment booths were set up in community events and community centres for children. In addition, we also approached children attending Hong Kong Children’s Hospital for conditions unrelated to WD such as those who attended a surgical outpatient clinic for assessment of circumcision. Only children who had no known history of liver, kidney, neurological, or trace element metabolism disorders were invited to join the study. Patient sample IDs were not known to anyone outside the research group.

Participants were requested to submit two spot urine samples and one 24-hour urine collection. They were instructed not to collect urine samples during acute illness. The two spot urine samples can be collected at any time on 2 different days. Spot urine samples were collected into acid-washed bottles which were specially prepared and validated for trace element analysis without contamination.

For 24-hour urine collection, participants were provided with detailed instructions and acid-washed collection containers. Completion of 24-hour collections was self-reported which was verified by measuring total volume and creatinine excretion.

### Screening Protocol and Cut-off Values

Previously developed cut-off values for 3 spot urine indices were evaluated based on our previous retrospective study (N. L. Tang et al. 2020):

- Spot urine copper concentration ≥0.5 µmol/L
- Copper to osmolality ratio (Copper/Osm) ≥0.00085 μmol/mOsm
- Copper to creatinine ratio (Copper/Cr) ≥0.1 µmol/mmol

Children whose spot urine sample exceeding any one cut-off value were called back for blood ceruloplasmin and copper as second-tier tests.

### Laboratory Methods

#### Measurement of Spot Urine Copper Concentrations

Urine copper was measured by inductively coupled plasma-mass spectrometry (ICP-MS) using an Agilent 7700 system. Urine samples were warmed to room temperature before dilution with pre-treatment standard solution. The samples were mixed with 3 internal standards, including yttrium, rhodium and iridium before loading into the ICPMS 7700 Analyser. Typically, a batch of analysis is performed together with standard calibrators and QC samples. In-house assay has a detection limit of 0.01 µmol/L for copper, which is at least one order of magnitude below the physiological urine concentration.

ICP-MS based copper measurement was performed in duplicate with appropriate quality control measures including: (1) Batch calibration with certified reference materials, (2) Analysis of internal quality control samples with each batch, (3) Participation in external quality assurance programs and (4) Contamination control through use of acid-washed collection containers and proper sample handling protocols.

#### Measurement of Other Urine Analytes

To generate meaningful excretion indices, spot urine concentrations of copper were normalised with either creatinine concentration or osmolality. Urine creatinine was measured by a modified Jaffe reaction on autoanalysers of Roche Diagnostics. Urine osmolality was measured by an analyser using the freezing-point method (Advanced Osmometers). In addition, spot urine protein was measured to identify patients with proteinuria. Patients with proteinuria (≥20 mg/mmol creatinine) due to glomerular disease may have albuminuria and ceruloplasminuria, which results in abnormally high copper excretion. Thus, spot urine copper as a screening test for WD is not valid in these subjects. Urine protein was measured by a turbidimetric method using ethylenediaminetetraacetic acid (EDTA) and benzethonium chloride.

#### Second-tier Testing

Children who exceeded screening cut-off values underwent second-tier testing including:

- plasma ceruloplasmin measurement by immunoturbidimetric assay
- plasma copper measurements by ICP-MS analyser.
- Mutation analysis of the *ATP7B* Gene. Genetic testing was performed for all call-back children after obtaining consent from their parent/guardians. DNA was extracted from peripheral blood samples for *ATP7B* gene sequencing and the entire coding sequence of *ATP7B* including exon-intron boundaries was analyzed for pathogenic variants.

#### Original study design and sample size considerations

Based on the estimated prevalence of WD in Hong Kong (∼1 in 5,000), this prospective study was not designed to pick up any WD cases but to confirm the reference range we developed in archival samples of pre-school children previously. Furthermore, we extended the age group to cover older school children so that future screening programs will have reference range data for a wider age group. Other screening programs can choose the appropriate age group for WD screening according to their local student health screening practices and policies. This sample size was sufficient to evaluate the reference intervals of urinary copper excretion indices and our previously reported cut-off values (Solberg 1993).

### Statistical Analysis

Descriptive statistics were used to characterise the study cohort and screening results. The distribution and performance of different spot urine copper indices were evaluated. Correlation between 24-hour urine excretion and spot urine indices was performed by regression through the origin approach (Eisenhauer 2003). The call-back rate was calculated as the percentage of children requiring second-tier testing. P-values <0.05 were considered statistically significant.

## RESULTS

### Characteristics of the Study Population

We recruited 225 healthy schoolchildren. 31 of them did not return any sample and one exceeded 11 years of age at the time of urine collection. At the end of the study, results of 193 children (86% participation rate) aged between 4-11 years were analysed (Figure 1).

**Figure 1.**
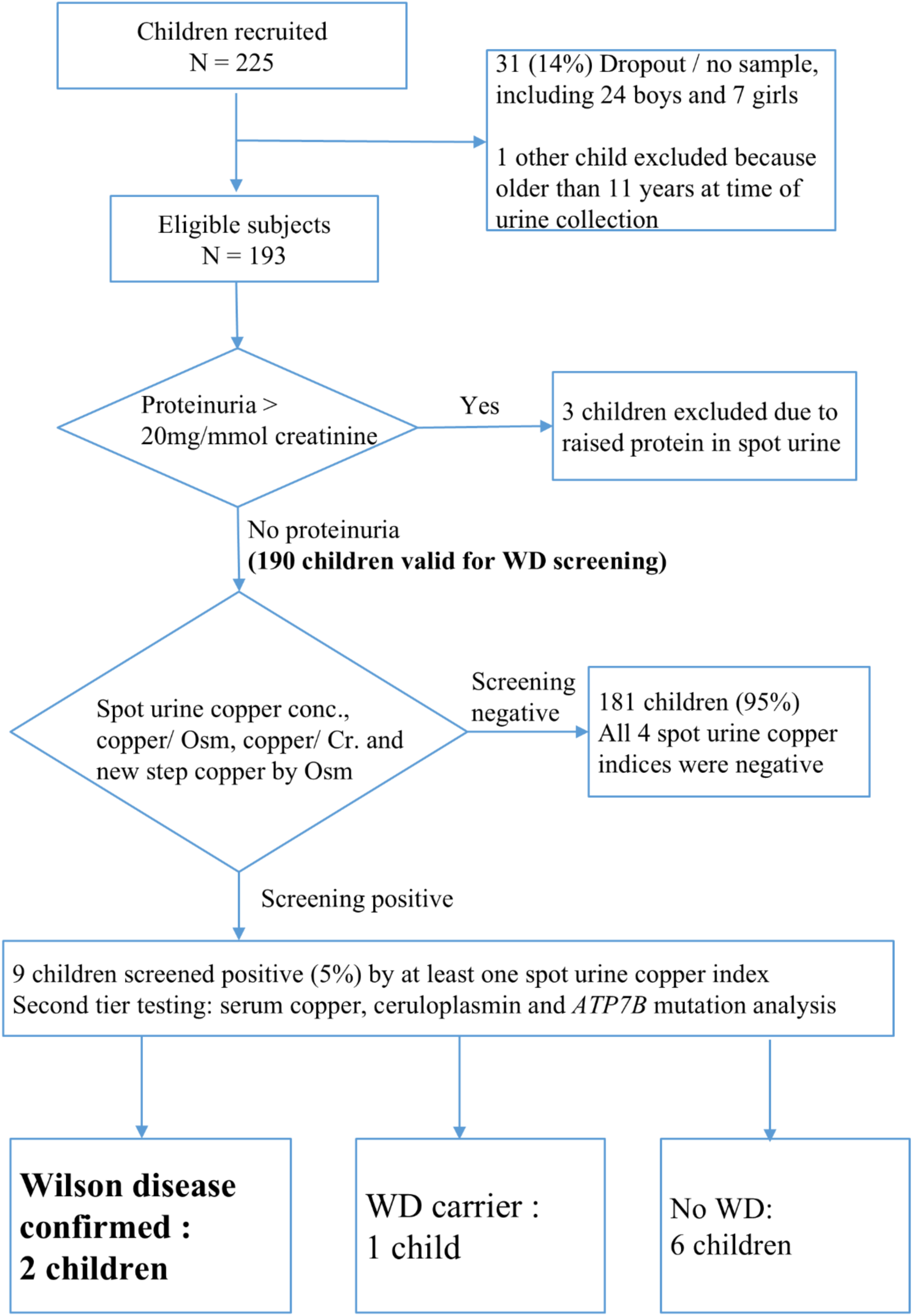
The details of participants and the Outcome of the WD screening are shown.

Among the 193 children, 176 (91%) returned the complete set of urine samples (2 spot urine and one 24-hour collection). The median age of participants was 7 years (range 4-11 years), with approximately 59% were in younger (4-7 years) and 41% were in older (8-11 years) age groups. There were no differences in demographic characteristics between 176 children who completed the study protocol and those who did not provide all 3 urine samples. 72% of participants were male. (Table 1)

**Table 1.** Age distribution of the 193 school children participants.

| Age (years) | Boys | Girls | Total number |
| --- | --- | --- | --- |
| **4-5** | 33 | 15 | 48 |
| **6-7** | 49 | 16 | 65 |
| **8-9** | 36 | 16 | 52 |
| **10-11** | 21 | 7 | 28 |
| **Total number** | 139 | 54 | 193 |
| **Median age (years)** | 7.0 | 7.0 | 7.0 |

### Characteristics of Urine copper indices

In total, there were 373 spot urine samples and 176 24-hour urine samples. As proteinuria could potentially lead to high urine copper excretion, spot urine protein was also measured. Three children had a spot urine protein above 20 mg/mmol creatinine and were referred to paediatric clinic for assessment. All were confirmed to have mild proteinuria of urine protein excretion less than 0.3 g per day. Their urine copper concentrations were 0.19, 0.41, 0.44, 0.46, 0.46 and 0.56 µmol/L which were significantly higher than the rest of the groups (p<0.05 on Mann-Whitney U test), so their data were excluded from the subsequent analysis.

### Spot urine copper excretion indices by age group

As our previous study was confined to pre-school children (N. L. Tang et al. 2020), we separated this cohort into 2 subgroups by age (Age 4 to 7 and Age 8 to 11) accordingly to better match the age group and allow comparison. There were 217 samples in the younger age group and 156 samples in the older age group. After excluding 12 spot urine samples collected from 3 children with proteinuria, the 2 WD cases and one heterozygote carrier subsequently diagnosed in the study, 361 spot urine samples were analysed for reference intervals and distribution of spot urine copper indices in healthy children (Table 2).

**Table 2.**
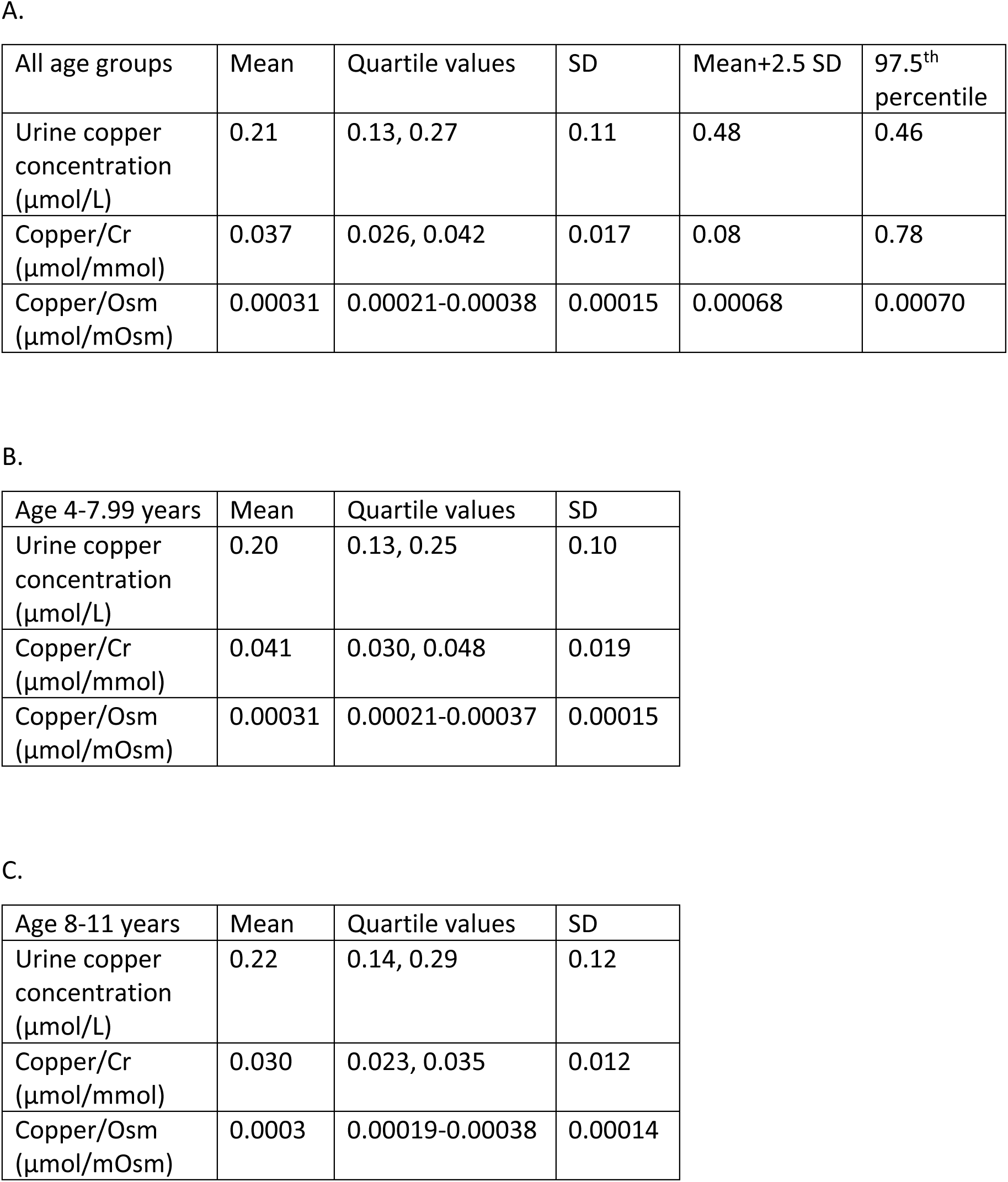
Distribution of spot urine copper excretion indices of healthy children (A: all age group; B: 4-7.99 years; C: 8-11 years), after exclusion of children with proteinuria, the 2 WD patients and one carrier (i.e. N= 361 spot urines from 187 children). Values of two potential screening cut-off points are shown (mean + 2.5 SD or the 97.5th percentile). SD: standard deviation.

| All age groups | Mean | Quartile values | SD | Mean+2.5 SD | 97.5<sup>th</sup> percentile |
| --- | --- | --- | --- | --- | --- |
| Urine copper concentration (μmol/L) | 0.21 | 0.13, 0.27 | 0.11 | 0.48 | 0.46 |
| Copper/Cr (μmol/mmol) | 0.037 | 0.026, 0.042 | 0.017 | 0.08 | 0.78 |
| Copper/Osm (μmol/mOsm) | 0.00031 | 0.00021-0.00038 | 0.00015 | 0.00068 | 0.00070 |

B.
| Age 4-7.99 years | Mean | Quartile values | SD |
| --- | --- | --- | --- |
| Urine copper concentration (μmol/L) | 0.20 | 0.13, 0.25 | 0.10 |
| Copper/Cr (μmol/mmol) | 0.041 | 0.030, 0.048 | 0.019 |
| Copper/Osm (μmol/mOsm) | 0.00031 | 0.00021-0.00037 | 0.00015 |

C.
| Age 8-11 years | Mean | Quartile values | SD |
| --- | --- | --- | --- |
| Urine copper concentration (μmol/L) | 0.22 | 0.14, 0.29 | 0.12 |
| Copper/Cr (μmol/mmol) | 0.030 | 0.023, 0.035 | 0.012 |
| Copper/Osm (μmol/mOsm) | 0.0003 | 0.00019-0.00038 | 0.00014 |

Mean urine copper concentration was 0.21 µmol/L (SD 0.11) (Table 2 and Suppl Figure 1). There was no age group difference in spot urine copper concentration. When urine copper was normalised to creatinine (Copper/Cr) or osmolality (Copper/Osm), the statistical distributions in this prospective cohort were similar to the previous report. The distribution of spot urine copper, Copper/Cr and Copper/Osm were compared between the 2 age subgroups and are shown in supplementary Figure 1 to 3, respectively. These indices do not have statistically significant differences between the 2 age-groups except Copper/Cr (Suppl Figure 2). Copper/Cr was lower in the elder children as they had a higher creatinine excretion. The mean plus 2.5 SD values were comparable to our previously study.

All 3 indices showed similar level of biological variation percentage (CVg%) of 52%, 46% and 48% for spot urine copper concentration, Copper/Cr and Copper/Osm, respectively. The results were consistent with our previous findings.

### Primary outcomes: Screening Results

Nine children were called back for results of their spot urine indices exceeding at least one cut-off value (Table 3). In addition, one child (Cu050) was called back because his 24-hour urine copper excretion was over the 95^th^ percentile of the group (0.49 μmol/day, or 31 μg/day). Among the whole cohort of 193 children, the call-back rate was about 5%. The distribution of positive screening results according to cut-off values of different indices is shown in Table 3. Details of their spot urine and 24-hour urine results are shown in supplementary tables 1 and 2.

**Table 3.** Number of participants exceeded the cut-off values of urine copper excretion indices. Both the single cut-off value and step cut-off criteria for urine copper concentration (shown in Bold font) can correctly identify both WD patients and also the carrier.

| Spot urine cut-off criteria (previously reported in(N. L. Tang et al. 2020)) | Number of children screened positive | % in 193 children | WD and carrier positive |
| --- | --- | --- | --- |
| **Urine copper $\geq 0.5 \mu\text{mol/L}$** | **7** | **3.7%** | **2/2, 1/1** |
| Copper/Osm $\geq 0.00085 \mu\text{mol/mOsm}$ | 6 | 3.2% | 2/2, 0/1 |
| Copper/Cr $\geq 0.1 \mu\text{mol/mmol}$ | 2 | 1.0% | 1/2, 0/1 |
| Modified step cut-off criteria (new cut-off values related to urine osmolality proposed) |  |  |  |
| **Urine copper $\geq 0.5 \mu\text{mol/L}$ if urine osmolality $\geq 600 \text{ mOsm/kg}$ or Urine copper $\geq 0.3 \mu\text{mol/L}$ if urine osmolality $< 600 \text{ mOsm/kg}$** | **9** | **4.7%** | **2/2, 1/1** |

As shown in Figure 2, the urine copper concentrations were higher in those samples with osmolality above 600 mOsm/kg. Therefore, the distribution of urine copper concentration were analysed separately by dividing the samples into two subgroups based on urine osmolality (urine osmolality≥600 mOsm/kg and urine osmolality<600 mOsm/kg). The mean (SD) of the 2 subgroups are shown in Supplementary Table 1 and formed the basis of the newly proposed spot urine copper concentration step cut-off criteria in Table 3.

**Figure 2.**
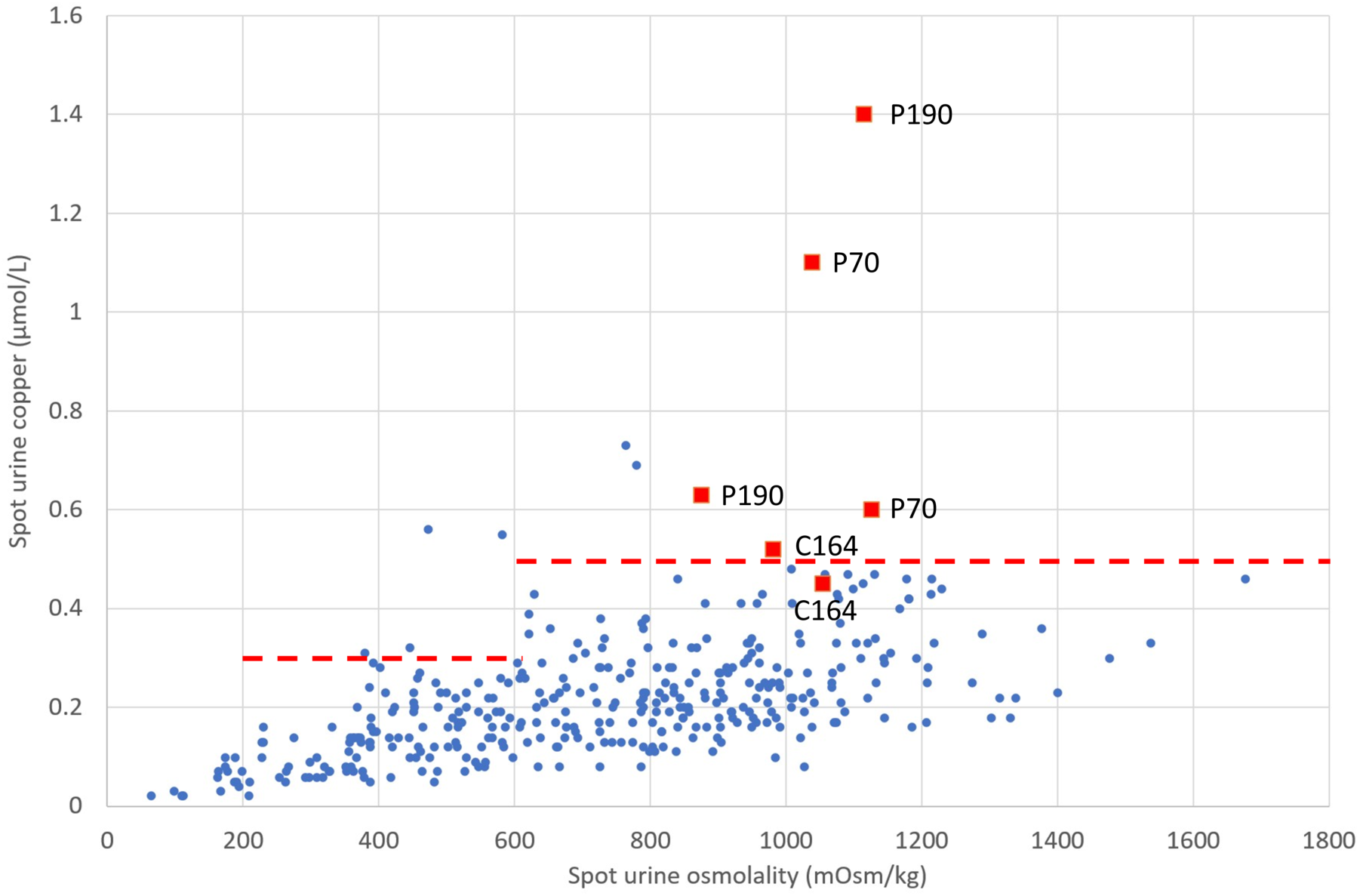
Spot urine copper concentration was related to spot urine osmolality. Based on the correlation between copper concentration and osmolality, a step cut-off levels were proposed (shown as dash lines). Samples results of two WD patients and the carrier picked up by this screening were labelled by squares. Samples of the two confirmed WD patients were shown as squares labelled with P70 and P190. The carrier were labelled C164.

### Second-tier Testing and Diagnosis

All 10 children recalled had serum ceruloplasmin and copper levels checked. Two children who are siblings had abnormally low ceruloplasmin levels. One of them also had elevated 24-hour urine copper excretion (0.77 μmol/day or 49 μg/day), which was above the diagnostic value of paediatric WD. On the other hand, the other sibling excreted a lower copper amount at 0.41 μmol/day (26μg/day) which were below the typical diagnostic values of WD used in children (0.62 μmol/day or 40 μg/day) (Nicastro et al. 2010; Socha et al. 2018).

### Performance of Screening Indices

Single spot urine copper concentration cut-off values (either as a single cut-off ≥0.5 umol/L or step cut-off related to urine osmolality described above) demonstrated the best screening performance, correctly identifying both confirmed WD cases. When analyzing the data retrospectively, both cut-off approaches (ie spot urine copper ≥0.5 μmol/L and step cut-off) had sufficient sensitivity to pick-up the 2 confirmed WD patients. And the specificity was also high at 95% (182/191 non-WD children).

### Genetic Confirmation

*ATP7B* gene mutation analysis was performed on the two siblings (P70 and P190) with biochemical findings consistent with WD. Both children were confirmed to have compound heterozygous mutations in the *ATP7B* gene – p.Thr935Met and p.Ile1148Thr – both are recognised pathogenic mutations in the local Chinese population (Table 4) (Cheng et al 2017, Li et al 2021).

**Table 4.** Details of cut-off exceeded in the 10 called back children and their mutation analysis results. In columns 2 to 5, counts of spot urine samples exceeded each spot urine index are shown. For detailed laboratory results, please refer to Supplementary Table 1.

| Subject ID | copper high $\geq 0.50$ | Copper/Cr high | Copper/Osm high | Step cut-off high | 24hr Ur Copper (umol/d) | Ceruloplasmin (mg/dl) | ATP7B Mutations |
| --- | --- | --- | --- | --- | --- | --- | --- |
| Cu099 | 1 | 0 | 1 | 1 | 0.2 | 23 | neg |
| Cu117 | 1 | 0 | 1 | 1 | 0.4 | 21 | neg |
| Cu015 | 1 | 0 | 1 | 1 | 0.15 | 31 | neg |
| Cu073 | 0 | 0 | 0 | 1 | 0.12 | 19 | neg |
| Cu066 | 0 | 0 | 0 | 1 | 0.1 | 27 | N.A. |
| Cu042 | 1 | 1 | 1 | 1 | 0.19 | 29 | neg |
| **2 WD patients and 1 carrier** |  |  |  |  |  |  |  |
| C164 | 1 | 0 | 0 | 1 | 0.12 | 22 | carrier |
| P070 | 2 | 0 | 1 | 2 | 0.41 | 6 | WD |
| P190 | 2 | 2 | 1 | 2 | 0.77 | <5 | WD |
| Called back because of higher 24-hr urine copper in the group |  |  |  |  |  |  |  |
| Cu050 | 0 | 0 | 0 | 0 | 0.49 | 24 | neg |

Both children were asymptomatic at the time of diagnosis, with normal liver function tests and no neurological symptoms. An MRI of the brain was performed and no abnormality was detected. Subsequently, both children started on zinc sulphate therapy (50 mg elemental zinc twice daily). At 6-month follow-up, both remained asymptomatic with normal liver function tests. Their parents were confirmed carriers of the mutations.

In addition, one carrier (C164) was identified who carried the mutation p.Arg919Gly which is also a locally prevalent mutation (Cheng et al 2017, Li et al 2021). The remaining recalled children were negative for pathogenic mutation after sequencing of the *ATP7B* gene.

### Additional Findings of children with proteinuria

During the screening process, the three children identified with incidental proteinuria were referred for appropriate nephrological evaluation and all were confirmed benign proteinuria at follow-up.

### Correlation between 24-hour Urine copper excretion and spot urine indices explains why spot urine can screen for WD

Among the 173 children (after exclusion of 2 WD and 1 carrier) who provided 24-hour urine samples, the mean (SD) of daily urine copper excretion was 0.12 (0.07) μmol/day. Inter-quartile range was 0.08 μmol/day to 0.16 μmol/day and the 97.5^th^ percentile was 0.32 μmol/day (∼21 μg/day). This data can be used as a reference interval for daily copper excretion in healthy children.

Both the amount of daily excretion and the concentration of copper in the 24-hour urine samples correlated with spot urine copper concentration. The correlation between 24-hour daily amount of copper excretion and average spot urine copper concentration was significant (R^2^=0.73, p<0.05). The correlation between the copper concentration in the 24-hour urine and average spot urine copper concentration was even stronger (R^2^= 0.83, p<0.05). (Figure 3)

**Figure 3.**
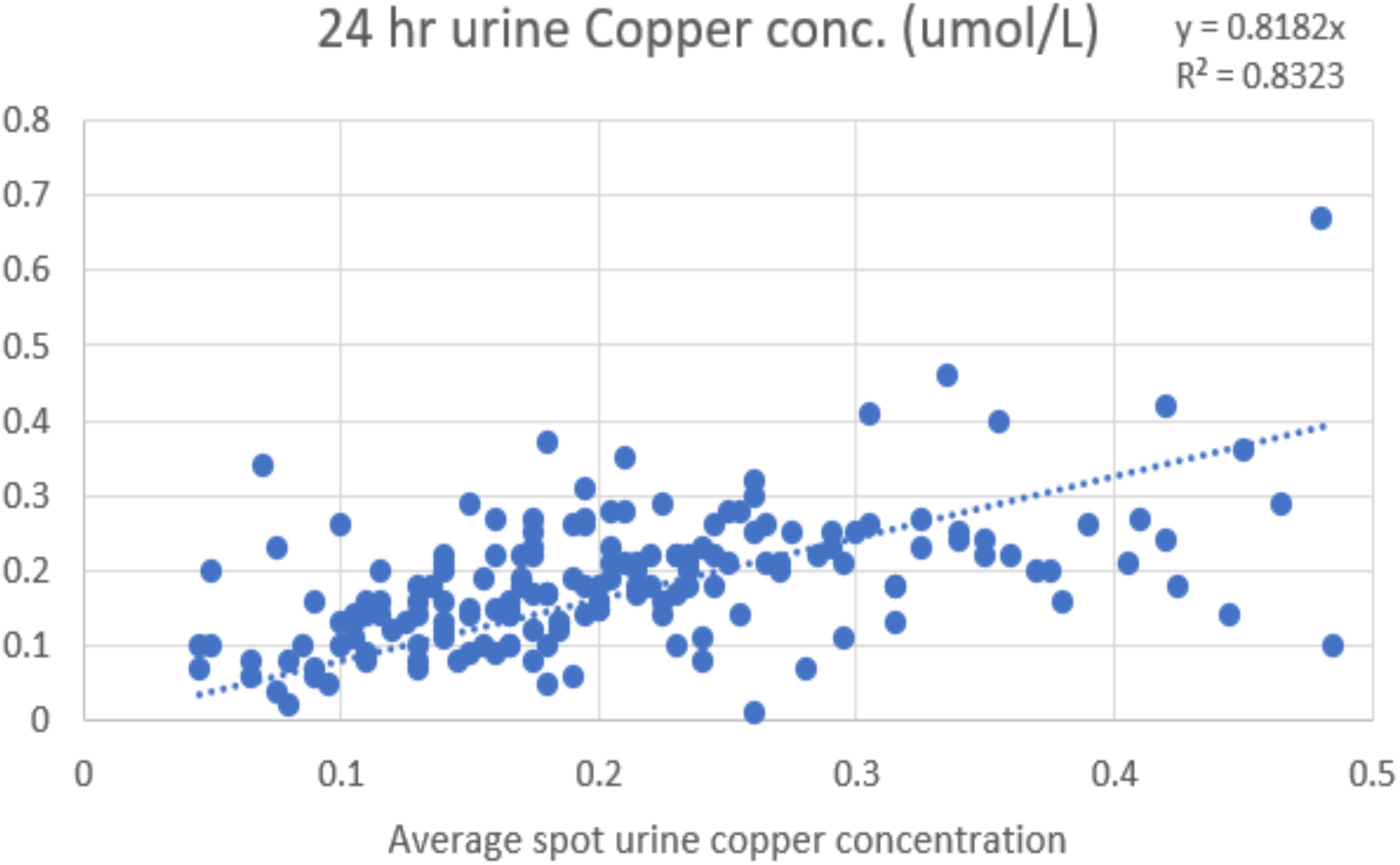
The correlation between urine copper concentration in the 24-hour urine collection and the average of copper concentration in 2 spot urines is statistically significant (P<0.01).

## DISCUSSION

This prospective validation study demonstrates that spot urine copper screening is useful in universal screening of WD in schoolchildren. Despite our small sample size of only approximately 200 children, we unexpectedly diagnosed 2 patients with pre-symptomatic WD, confirming the clinical utility of our screening approach. The spot urine copper concentration (both as a single cut-off or a step cut-off related to urine osmolality) was shown to be the most reliable screening cut-off, with excellent sensitivity and high specificity.

### Comparison with other screening approaches

Previous attempts at WD screening faced numerous challenges, e.g. measurement of ceruloplasmin in NBS reported low success rates (reviewed in (Hahn 2014)). Recently, a newly developed immune-SRM method using NBS dried blood spots to quantitate ATP7B peptides showed promising results (Jung et al. 2017; Collins et al. 2021; Klippel et al. 2025). Here, our spot urine screening approach added to the repertoire of biomarkers that are potentially useful in universal screening of WD. The spot urine approach differs fundamentally by: (1) targeting school-age children when copper accumulation becomes detectable but before symptom onset, (2) being non-invasive, using spot urine copper rather than blood-based markers, and (3) not requiring specialised technology like the immuno-SRM method while just employing more widely available ICP-MS technology and machines.

### Clinical Implications

The successful identification of pre-symptomatic WD cases by spot urine biomarkers in this small pilot study is encouraging. Generally, organ damage in WD first presents with liver dysfunction usually in teenagers and neurological symptoms may follow a decade later (Loudianos et al. 2024; Socha et al. 2018). Therefore, there is a 10-year window period for screening to pick up pre-symptomatic WD. The design of our urine screening targeting primary school children (4-11 years) fits into this critical pre-symptomatic period. Recent studies confirmed that monotherapy with zinc was highly effective and there are cases whose clinical symptoms of copper toxicity did not develop after early treatment (Chanpong and Dhawan 2022; Loudianos et al. 2024). This represents a paradigm shift from reactive treatment of WD patients with symptomatic disease to proactive prevention of organ damage by early treatment in the pre-symptomatic phase.

### Screening Feasibility and Acceptability

Our study demonstrates excellent acceptance and feasibility with an 86% participation rate for spot urine collection. The spot urine collection was acceptable to most children, as it is non-invasive and requires minimal time commitment. In the study setting, 24-hour urine was also collected. 24-hour urine collection will be unnecessary in the practice of universal screening, as we have shown here that spot urine copper indices correlated strongly with 24-hour urine copper excretion, which supports the use of spot urine in screening of WD. Furthermore, spot urine collection also addressed the drawbacks of 24-hour urine collection in paediatric populations. The call-back rate of approximately 5% is reasonable compared to other childhood screening programs and suggests that screening implementation would not overwhelm healthcare resources.

The use of ICP-MS technology for the measurement of trace elements in biological samples represents a major advance in the last decades, and this has been widely accessible in pathology laboratories worldwide. With its detection limits of 0.01 µmol/L for copper, well below physiological ranges and the cut-off value (0.5 µmol/L), this technology enables accurate measurement of trace element concentrations and historical measurement problems associated with flame photometry were greatly improved. However, the level of 0.5 µmol/L has not been included in most external quality control programs at the moment, there is a need for future external quality control programs to provide QC samples covering this low range.

### Universal Screening Criteria are met

Our screening approach fulfils the modern screening criteria (Andermann et al. 2008):

1. WD is an important health problem with an incidence in Hong Kong as high as 1 in 5,400
2. Zinc monotherapy represents an accepted and highly effective treatment
3. Pre-symptomatic WD is now a recognised early phase of disease
4. Our study validates spot urine copper as a suitable screening test
5. The test is acceptable to the population and economically viable
6. The 10-year period in the natural history of WD from pre-symptomatic to symptomatic disease provides the window for universal screening

Based on our successful pilot study, we propose several directions for future research:

1. Multi-centre validation studies involving more children of various ethnic backgrounds to confirm and refine the cut-off values of spot urine copper indices
2. Longitudinal follow-up of pre-symptomatic or screen-detected cases to document long-term outcomes
3. Explore how to integrate with local school health programs for implementation (Figure 4)

**Figure 4.**
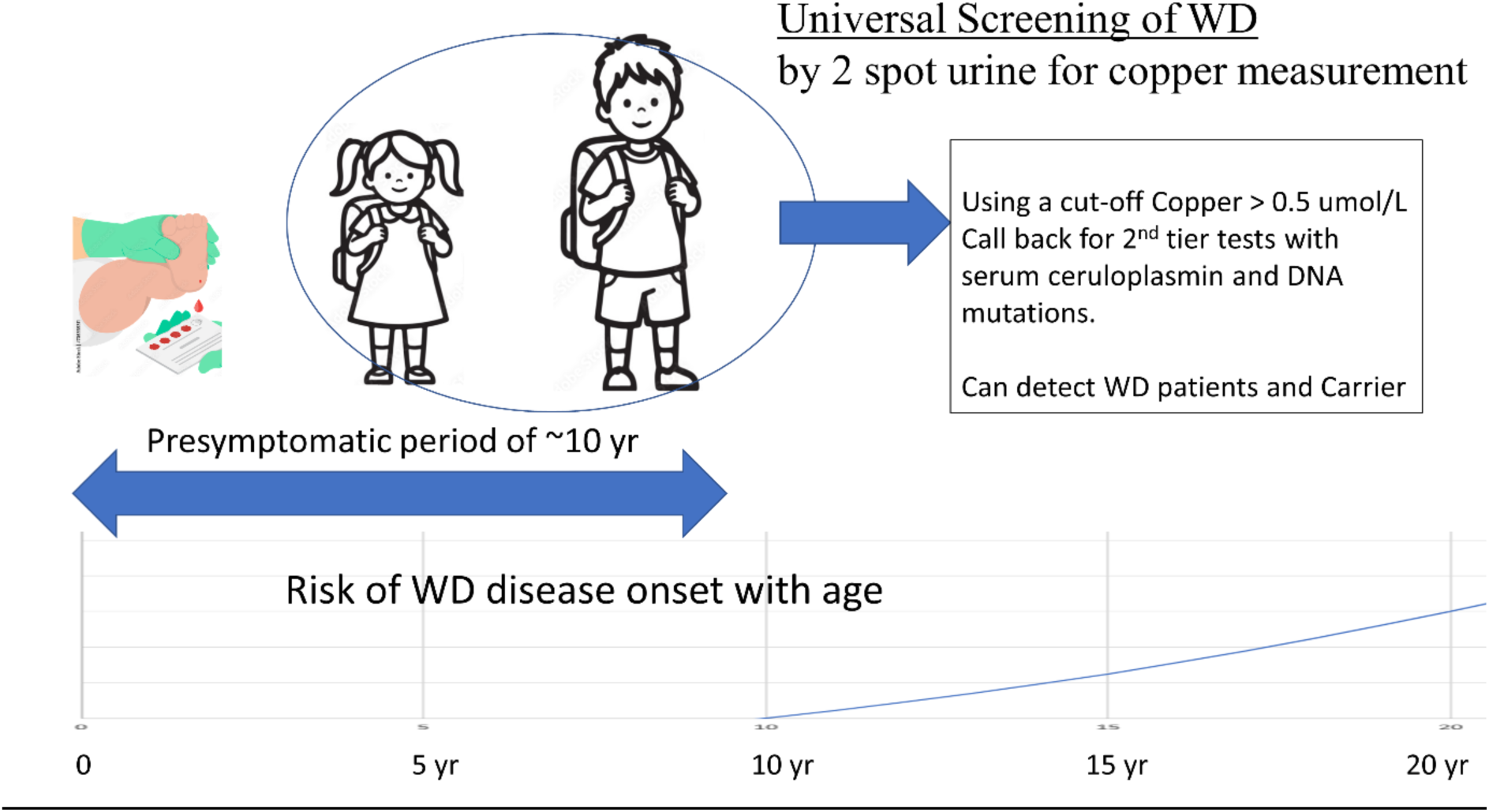
Spot urine screening approach targeting school children provides a new approach to universal screening of WD in addition to NBS. Both approaches can identify per-symptomatic WD patients before disease onset.

Our findings suggest that universal screening of school-age children using spot urine copper measurement could fundamentally alter the landscape of WD management. Countries with high WD prevalence should consider implementation of similar screening programs. The relatively simple technological requirements and acceptable cost profile make this approach feasible even in developing countries or settings under limited resources.

### Limitations

First, our sample size was limited to approximately 200 children, which may affect the generalizability of our findings. And the detection of two siblings with WD represents a family cluster that inflates the apparent prevalence in our cohort.

Second, we did not sequence the *ATP7B* gene for all subjects in this prospective cohort, so we cannot be certain that the two siblings were the only WD patients in the cohort. However, given the prevalence of WD is between 1 in 5,000 and 1 in 17,000 in Chinese (J. Wang and Abuduxikuer 2019; Mak et al. 2008), we did not expect to have any sampled WD patients in the original study plan. Therefore, it is a fair assumption that there are only 2 WD patients in the screening cohort.

### Conclusion

This prospective study provides compelling evidence that spot urine copper screening can effectively identify pre-symptomatic WD in school-age children. The unanticipated success in detecting 2 WD patients together with the carrier highlights the effectiveness and potential impact of urine screening for WD. Given the paucity of biomarkers useful for universal screening of WD, the spot urine screening approach reported here represents a valuable addition to the limited repertoire of WD screening methods (Figure 4). Using a simple cut-off ≥0.5 µmol/L or a step cut-off as the primary screening criteria, we have demonstrated a practical, acceptable, and effective approach to early WD detection that could fundamentally improve patient outcomes by enabling pre-symptomatic treatment and preventing irreversible complications.

## Data Availability

All data produced in the present study are available upon reasonable request to the authors

## Acknowledgement

We thank Dr. Tsz-Ki Kwan for valuable suggestions and Jenkin Tang for assistance in language editing and proofreading. Funding support by Health and Medical Research Fund: PR-HKCH-10. Grammarly was used for the correction of spelling and grammar.

## Authors contribution

Conceptualization: NT, AMK, JH.

Funding acquisition and project supervision: AMK, JH, KB, AL, CF, NT.

Subject recruitment: AMK, FY, YT, AL, CF, NT.

Urine screening tests: IC, NC, XW, NT.

Clinical care and call back: AMK, CF.

Mutation analysis and genetic counseling: TCC, LH, THC, MY, CM.

Writing of original draft: NT, AMK, IC, AL.

All authors involved in editing and revision of the manuscript and approve the final version.

## Conflict of interest

None

## Supplementary figures

**Supplementary Figure 1.**
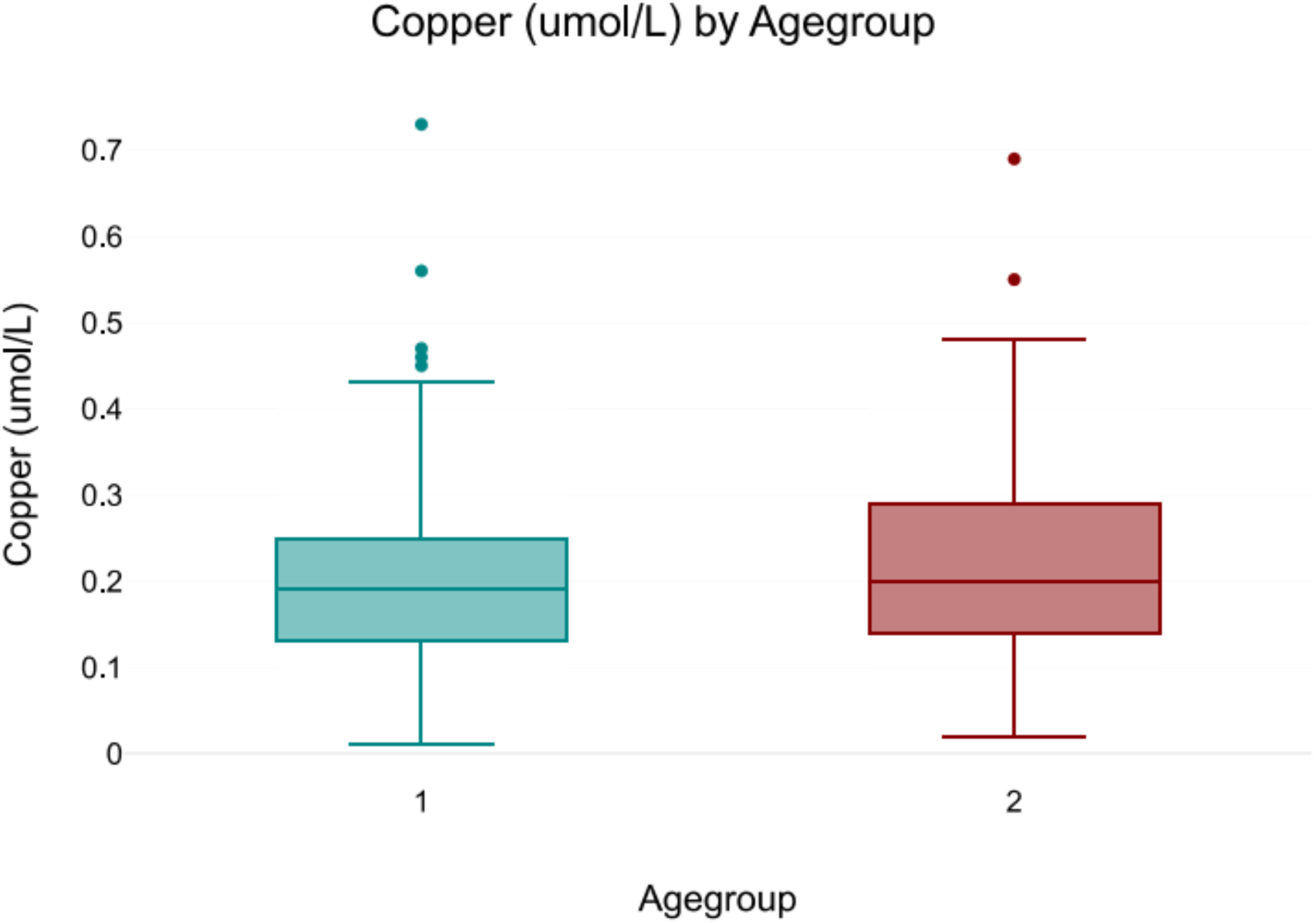
Distribution box plots of spot urine copper concentration of the 2 age groups. (1: 4 to 7.99 years old, 2: 8 to 11 years old). There was no statistical difference between the 2 groups (Mann-Whitney U test).

**Supplementary Figure 2.**
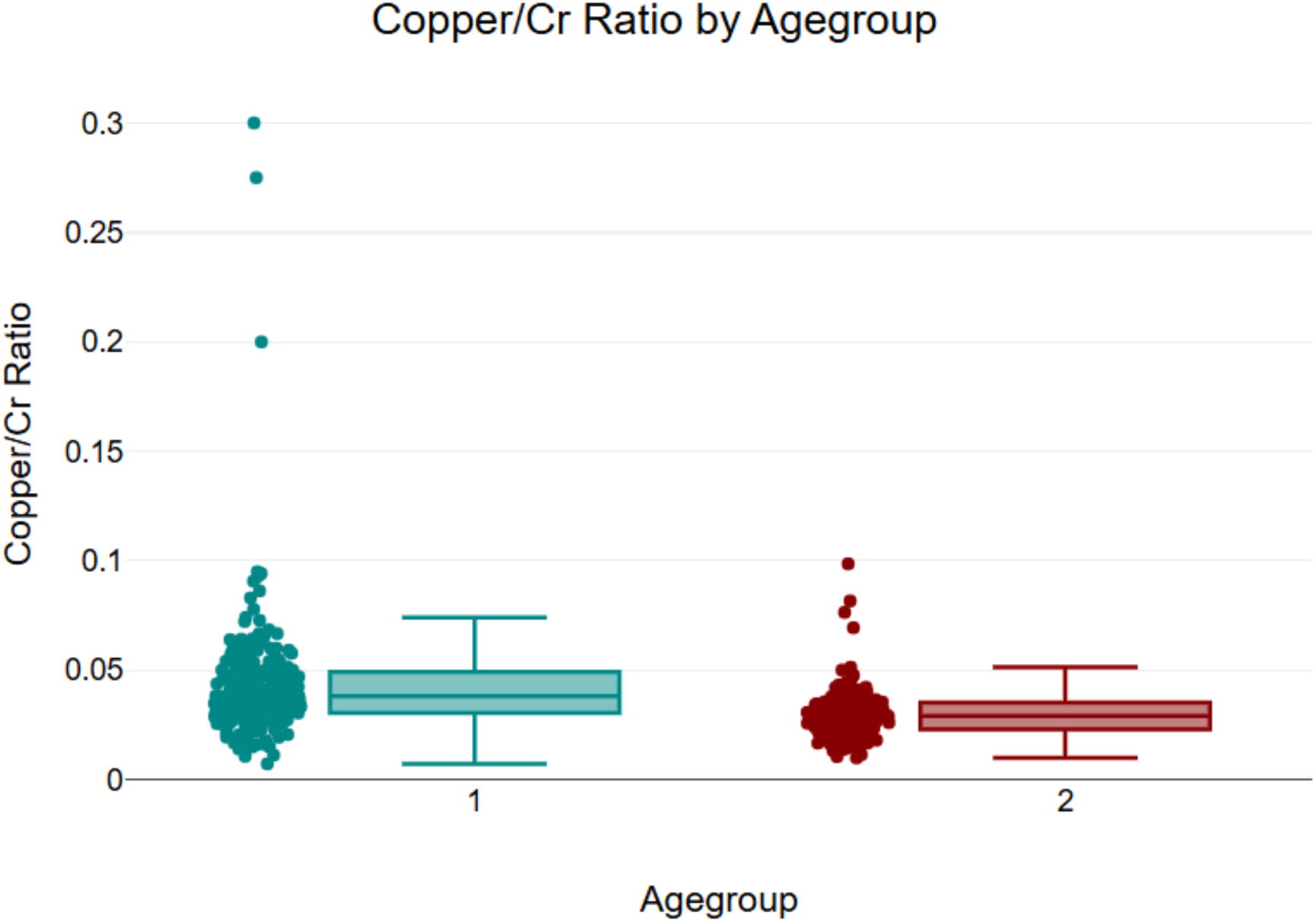
Distribution box plots of spot urine copper to Creatinine (Copper/Cr) ratio of the 2 age groups. (1: 4 to 7.99 years old, 2: 8 to 11 years old). There was a statistical difference between the 2 groups (p<0.001, Mann-Whitney U test).

**Supplementary Figure 3.**
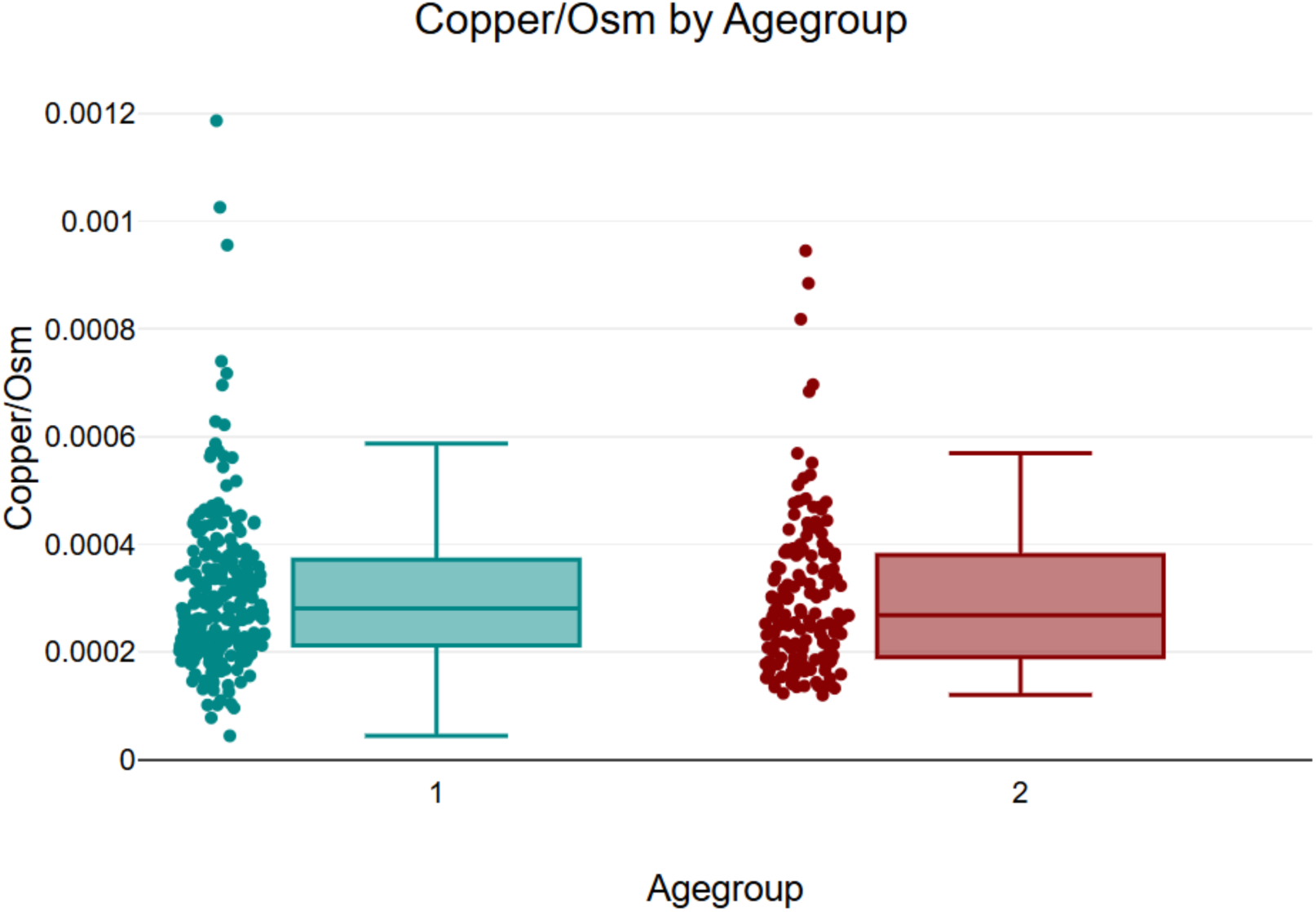
Distribution box plots of spot urine copper to osmolality (Copper/Osm) ratio of the 2 age groups. (1: 4 to 7.99 years old, 2: 8 to 11 years old). There was no statistical difference between the 2 groups (Mann-Whitney U test).

**Supplementary Figure 4.**
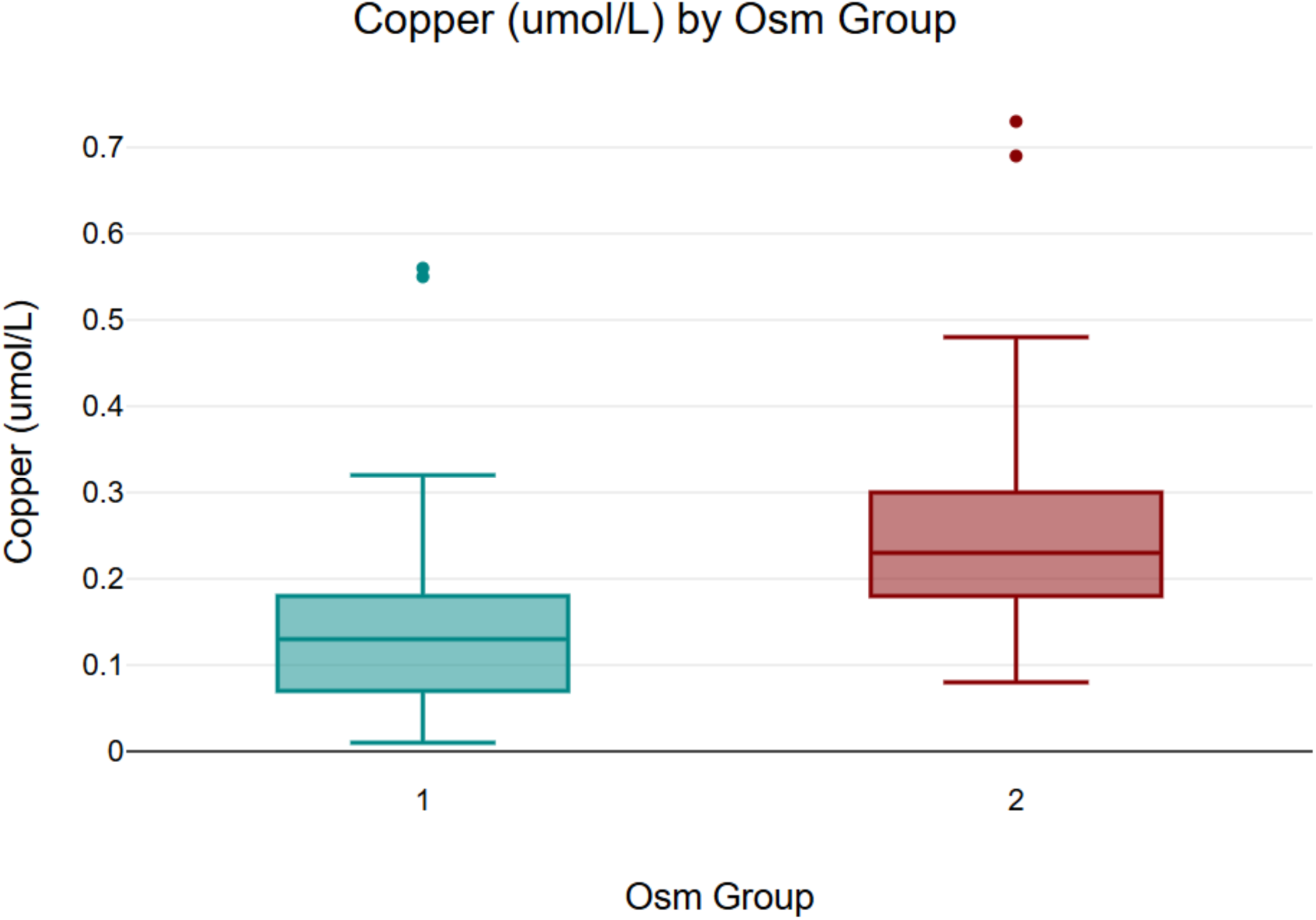
Distribution of urine copper related to urine osmolality. Osm groups are defined as 1: <600 mOsm/L and 2. >=600 mOsm/L. The cutoff value of urine copper for low and high urine osmolality groups are 0.3 umol/L and 0.5 umol/L, respectively. These values form the step cut-off values proposed here.

## Supplementary Tables

**Supplementary Table 1.**
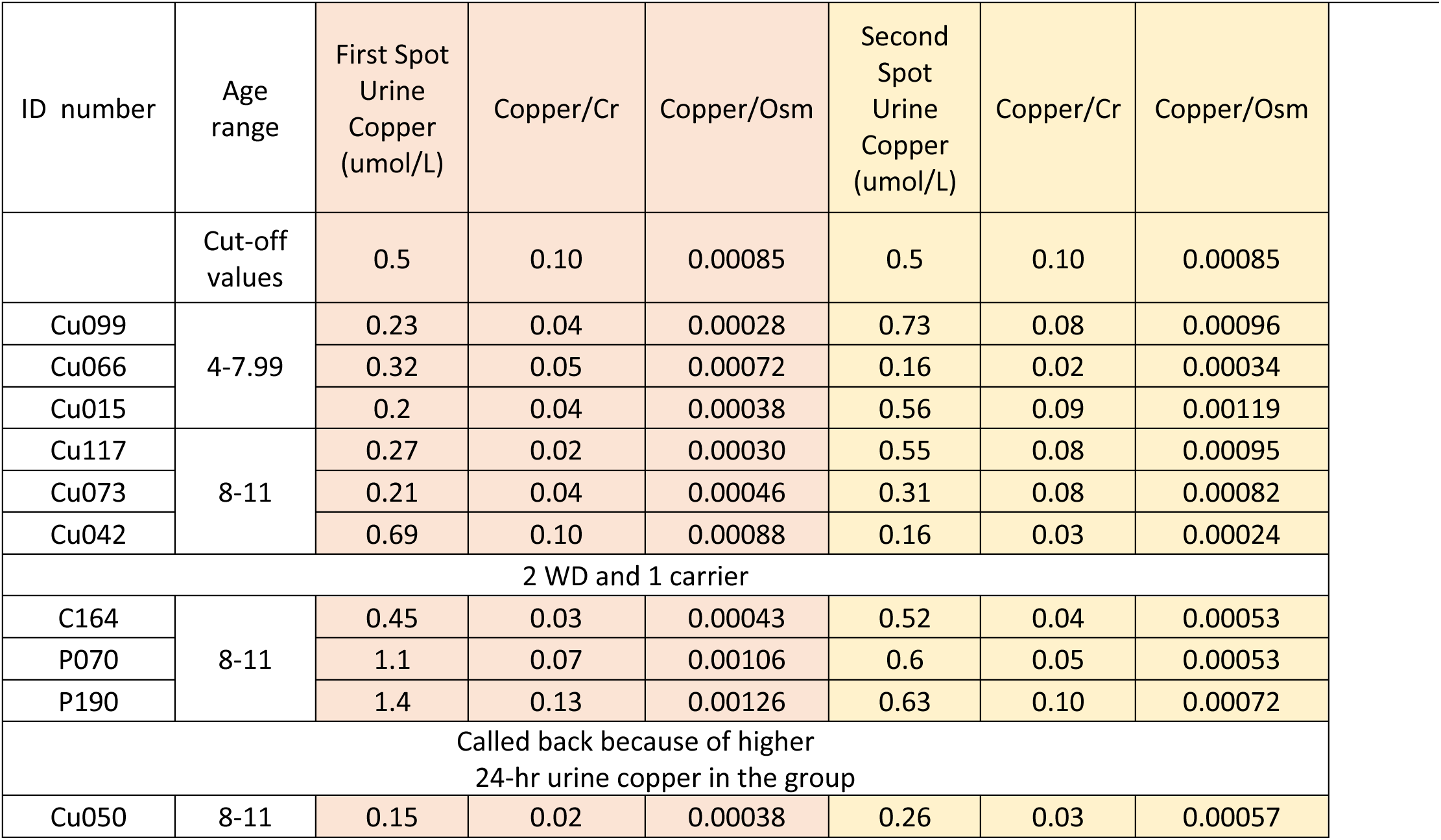
Details of spot urine results of 11 children called back for second tiers testing. The 2 WD patients are labelled P070 and P190. The carrier is C164. Cu050 was called backed because his 24 hour urine copper excretion was over 95^th^ percentile of the group. Values exceeding cut-off levels are marked in red font color. #Another boy (not shown in tables) was also called back whose spot urine copper was 0.48 umol/L, just below the cutoff, and his second tier investigations were negative.

| ID number | Age range | First Spot Urine Copper (umol/L) | Copper/Cr | Copper/Osm | Second Spot Urine Copper (umol/L) | Copper/Cr | Copper/Osm |
| --- | --- | --- | --- | --- | --- | --- | --- |
|  | Cut-off values | 0.5 | 0.10 | 0.00085 | 0.5 | 0.10 | 0.00085 |
| Cu099 | 4-7.99 | 0.23 | 0.04 | 0.00028 | 0.73 | 0.08 | 0.00096 |
| Cu066 |  | 0.32 | 0.05 | 0.00072 | 0.16 | 0.02 | 0.00034 |
| Cu015 |  | 0.2 | 0.04 | 0.00038 | 0.56 | 0.09 | 0.00119 |
| Cu117 | 8-11 | 0.27 | 0.02 | 0.00030 | 0.55 | 0.08 | 0.00095 |
| Cu073 |  | 0.21 | 0.04 | 0.00046 | 0.31 | 0.08 | 0.00082 |
| Cu042 |  | 0.69 | 0.10 | 0.00088 | 0.16 | 0.03 | 0.00024 |
| 2 WD and 1 carrier |  |  |  |  |  |  |  |
| C164 | 8-11 | 0.45 | 0.03 | 0.00043 | 0.52 | 0.04 | 0.00053 |
| P070 |  | 1.1 | 0.07 | 0.00106 | 0.6 | 0.05 | 0.00053 |
| P190 |  | 1.4 | 0.13 | 0.00126 | 0.63 | 0.10 | 0.00072 |
| Called back because of higher 24-hr urine copper in the group |  |  |  |  |  |  |  |
| Cu050 | 8-11 | 0.15 | 0.02 | 0.00038 | 0.26 | 0.03 | 0.00057 |

## Notes

### Competing Interest Statement

The authors have declared no competing interest.

### Funding Statement

This study was funded by Health and Medical Research Fund:
PR-HKCH-10

### Author Declarations

The study has been given ethical approval by the Institutional Review Board of the Hong Kong Children Hospital and the Joint CUHK-NTEC Clinical Research Ethics Committee.

